# COVID-19 vaccine hesitancy January-May 2021 among 18-64 year old US adults by employment and occupation

**DOI:** 10.1101/2021.04.20.21255821

**Authors:** Wendy C. King, Max Rubinstein, Alex Reinhart, Robin J. Mejia

## Abstract

COVID-19 vaccine hesitancy threatens pandemic control efforts. We evaluated vaccine hesitancy in the US by employment status and occupation category during the COVID-19 vaccine rollout. US adults 18-64 years completed an online COVID-19 survey 3,179,174 times from January 6-May 19, 2021. Data was aggregated by month. Survey weights matched the sample to the US population age, gender, and state profile. Weighted percentages and 95% confidence intervals (95%CI) were calculated. Changes in vaccine hesitancy from January-May varied widely by employment status (e.g., -7.8% [95%CI, -8.2--7.5] among those working outside the home, a 26.6% decrease; -13.3% [95%CI, -13.7--13.0] among those not working for pay, a 44.9% decrease), and occupation category (e.g., -15.9% [95%CI, -17.7--14.2] in production, a 39.3% decrease; -1.4% [95%CI, -3.8--1.0] in construction/extraction, a 3.0% decrease). April 20-May 19, 2021, vaccine hesitancy ranged from 7.3% (95%CI, 6.7-7.8) in computer/mathematical professions to 45.2% (95%CI, 43.2-46.8) in construction/extraction. Hesitancy was 9.0% (95%CI, 8.6-9.3) among educators and 14.5% (95%CI, 14.0-15.0) among healthcare practitioners/technicians. While the prevalence of reasons for hesitancy differed by occupation, over half of employed hesitant participants reported concern about side effects (51.7%) and not trusting COVID-19 vaccines (51.3%), whereas only 15.0% didn’t like vaccines in general. Over a third didn’t believe they needed the vaccine, didn’t trust the government, and/or were waiting to see if it was safe. In this massive national survey of adults 18-64 years, vaccine hesitancy varied widely by occupation. Reasons for hesitancy indicate messaging about safety and addressing trust are paramount.

## 1. Introduction

The development of highly efficacious COVID-19 vaccines in less than one year is a major medical accomplishment of the last century. However, vaccine hesitancy (i.e., a refusal or reluctance to be vaccinated) has slowed projected uptake^1^ and remains a barrier COVID-19 pandemic control.^2^ A longitudinal study of US adults that collected data through the approval and launch of three COVID-19 vaccines reported a decrease in COVID-19 vaccine hesitancy from 46.0% in October 2020 to 35.2% in March 2021^3^. Still, a greater reduction in vaccine hesitancy is needed to meet uptake goals of 70%-90%^1^.

Adults ≥60 years had a larger decrease in COVID-19 vaccine hesitancy versus younger adults October 2020-March 2021^3^, and consistent with previous reports^4,5,6^, had lower hesitancy at a given time point compared to younger adults. While younger versus older adults are less likely to be hospitalized or die from COVID-19^7^, vaccine hesitancy among working-age adults may contribute to workplace outbreaks and spread of infection between workers and customers, healthcare workers and patients, and educators and students, all serious public health threats^8,9^.

Age, sex, gender, race/ethnicity, education level, and living in an urban versus rural county are known correlates of COVID-19 vaccine hesitancy^3–6,10^. However, very few studies have evaluated COVID-19 vaccine hesitancy by employment status; those that did had small samples and were conducted in June 2020 about a then-future vaccine^4,5^, and we know of none comparing COVID-19 vaccine hesitancy by occupation. Elucidating the prevalence of vaccine hesitancy in the US workforce, and in particular, by occupation, is important for understanding risk of transmission and outbreaks in various job settings. Further, understanding why individuals are hesitant and if reasons vary by occupation is important for developing effective campaigns to increase vaccination uptake.

Among a massive sample of working-age (18-64 year old) US adults, we report COVID-19 vaccine hesitancy by month, January 6 through May 19, 2021, and evaluate time trends by employment status and occupation category. For the last 30 days, we report COVID-19 vaccination history and prevalence of COVID-19 vaccine hesitancy by occupation category, and the relative association between occupation category with hesitancy, with and without adjustment for demographics. Given healthcare workers and educators pose transmission risk to vulnerable populations (i.e., to patients, and children <12 years, who are not yet eligible for vaccination, respectively), we also evaluate hesitancy by profession within each of these occupations. Finally, we identify the most common reasons for COVID-19 vaccine hesitancy among the workforce and by occupation category.

## 2. Materials and Methods

### Sampling and weighting

Since April, 2020, the Delphi Group at Carnegie Mellon University (CMU) has been conducting an ongoing national survey, COVID-19 Trends and Impact Survey^11^, in collaboration with the Facebook Data for Good group. Each month the survey is offered to a random sample, stratified by geographic region, of ≈100 million US residents from the Facebook Active User Base who use one of the supported languages (English [American and British], Spanish [Spain and Latin American], French, Brazilian Portuguese, Vietnamese, and simplified Chinese). The offer to participate is shown with a link to the survey at the top of users’ Facebook News Feed to yield ≈1.1 million responders, which allows for evaluation of local trends.

When individuals click through the link, an anonymized unique identifier is generated. CMU returns the unique IDs to Facebook, which creates weights that account for the sampling design and non-response; these weights are then post-stratified to match the US general population by age, gender, and state^12^. This design safeguards respondent privacy by ensuring that researchers at CMU do not receive an identifying information about respondents and Facebook does not see survey microdata. The CMU Institutional Review Board approved the survey protocol and instrument (STUDY2020_00000162).

### Study sample

Facebook users may be offered the survey from once a month to once every six months, depending on their geographic strata. To show trends over time in vaccine hesitancy, we used data from January 6 to May 19, 2021 (a period in which the same version of the vaccine uptake and intent questions were offered to all potential respondents) aggregated by month. While it is possible there are repeat respondents across months, respondents cannot be linked longitudinally, so data was treated as repeat cross-sectional surveys. Only data from the last 30 days (April 20-May 19) was used in the cross-sectional analysis of vaccine uptake and hesitancy by occupation category/profession and reasons for hesitancy, avoiding repeat respondents and focusing on the most current data.

April 20-May 19, 2021, 104,760,491 Facebook users were offered the survey, of whom 904,022 completed at least two survey questions. Respondents were excluded if they were 65 or older (n=224,197), did not report their age (n=153,665,) or did not answer the vaccine acceptance question (n=351), leaving 525,809 participants. Applying the same criteria, the January-May monthly samples for time trends had 791,716; 710,529; 732,308; 631,621; and 313,000 participants, respectively; study flow by months is reported in supplemental **sTable 1**.

### Measures

The survey questions and response sets utilized in this report and a listing of professions by occupation category, based on the Bureau of Labor Statistics Standard Occupational Codes^13^, are provided in an **appendix** (supplemental material). The gender question was developed for this survey; other demographic questions were adapted from existing surveys: race and ethnicity match the 2020 Census definitions^14^, education categories were adapted from the American Community Survey^15^, age categories match the 10-year blocks reported by the ACS^16^. Participant’s self-reported home zip code was used to determine the urban-rural level of their metropolitan statistical area classification^17^. Vaccination questions were adapted from CDC-sponsored questions developed for two household panel surveys^18^ and shared with us prior to launch. The answer set for reasons for vaccine hesitancy, which appears to be a distinct phenomenon from general vaccine hesitancy, was expanded through a review of media reports and brainstorming sessions among survey methodologists.

For this analysis, participants were categorized as vaccine hesitant if they answered that they would “probably not” or “definitely not” choose to get vaccinated if offered a vaccine to prevent COVID-19 today (versus probably or definitely would choose to get vaccinated or were vaccinated), and as strongly hesitant if they answered “definitely not.” Already vaccinated individuals were included in the vaccine accepting category to ensure a consistent study population, as access to vaccinations varied by employment category, state, and month in the studied timeframe. Participants were categorized by employment status in the past 4 weeks (employed for pay, work outside the home; employed for pay, work at home; not employed for pay), and if employed, by occupation category and profession.

### Statistical analysis

All estimates were generated using survey weights^19.^ Percentage vaccine hesitant was calculated by month, overall, by employment status, and by occupation category. The difference in hesitancy from January to May was calculated as the May value minus the January value. The percent change was calculated as the difference divided by the January value. Percentages of employment status categories were also calculated by month to understand temporal trends in employment.

Among the final 30-day sample, percentages for worked outside the home, history of COVID-19 vaccination, and vaccine hesitancy (strong and total) were calculated among employed participants, by occupation categories, and by profession among health care practitioners/technicians, healthcare support and educators due their contact with vulnerable populations (i.e., patients, who may be high-risk for poor COVID-19 outcomes, or children, who may not yet be eligible for vaccination). Additionally, risk ratios (RR) for vaccine hesitancy by occupation category were calculated using Poisson regression. Adjusted RR were also calculated controlling for gender, age, race/ethnicity, education level, and urban-rural classification.

Finally, percentages for reasons for hesitancy were calculated among all employed vaccine hesitant participants; among healthcare practitioners/technicians, healthcare support, and educators; among the 5 occupation categories with the highest hesitancy prevalence, and among an additional 5 occupation categories with high-density indoor workspaces or significant client contact. For all parameters, 95% confidence intervals (95%CI) were calculated using robust standard errors.^8^ Analyses were conducted in R (Version 4.0.2, R Core Team, Vienna, Austria).

## 3. Results

### Participant characteristics

Final month (April 20-May 19) participants (N=732,308) had a median age range of 35-44 years; 45.5% were male, 52.0% female, 1.3% non-binary, and 1.2% self-described gender; 16.7% were Hispanic, 68.8% White, 6.5% Black, 3.6% Asian, 0.9% Native American, 0.3% Pacific Islander, and 3.4% Multi-racial; 23.2% had ≤high school education, 40.7% had ≥four-year college; 13.4% lived in a non-core or micropolitan area, 50.4% lived in a large central or fringe metro area. Two-thirds (66.1%) worked for pay; half (50.6%) worked outside the home. Demographics were similar across all months (data not shown), including employment status. Compared to January, in May: 1.7% more participants reported working outside the home, while 1.2% fewer reported working at home, and 0.4% fewer reported not working for pay (**eTable 2**).

### January-May time trends

As shown in **Figure 1 panel A** and reported in **eTable2**, vaccine hesitancy decreased 9.5 (95%CI, 9.3-9.7) percentage points, a 34.5% (95%CI, 35.2-33.8) decrease, from January (27.4% [95%CI, 27.3-27.6]) to May (18.0% [95%CI, 17.8-18.1]). There was a smaller relative decrease among those who worked outside the home (7.8 percentage points; a 26.6% decrease) versus those who worked from home (6.4 percentage points; a 42.4% decrease) or did not work for pay (13.3 percentage points; a 44.9% decrease).

**Figure 1.**
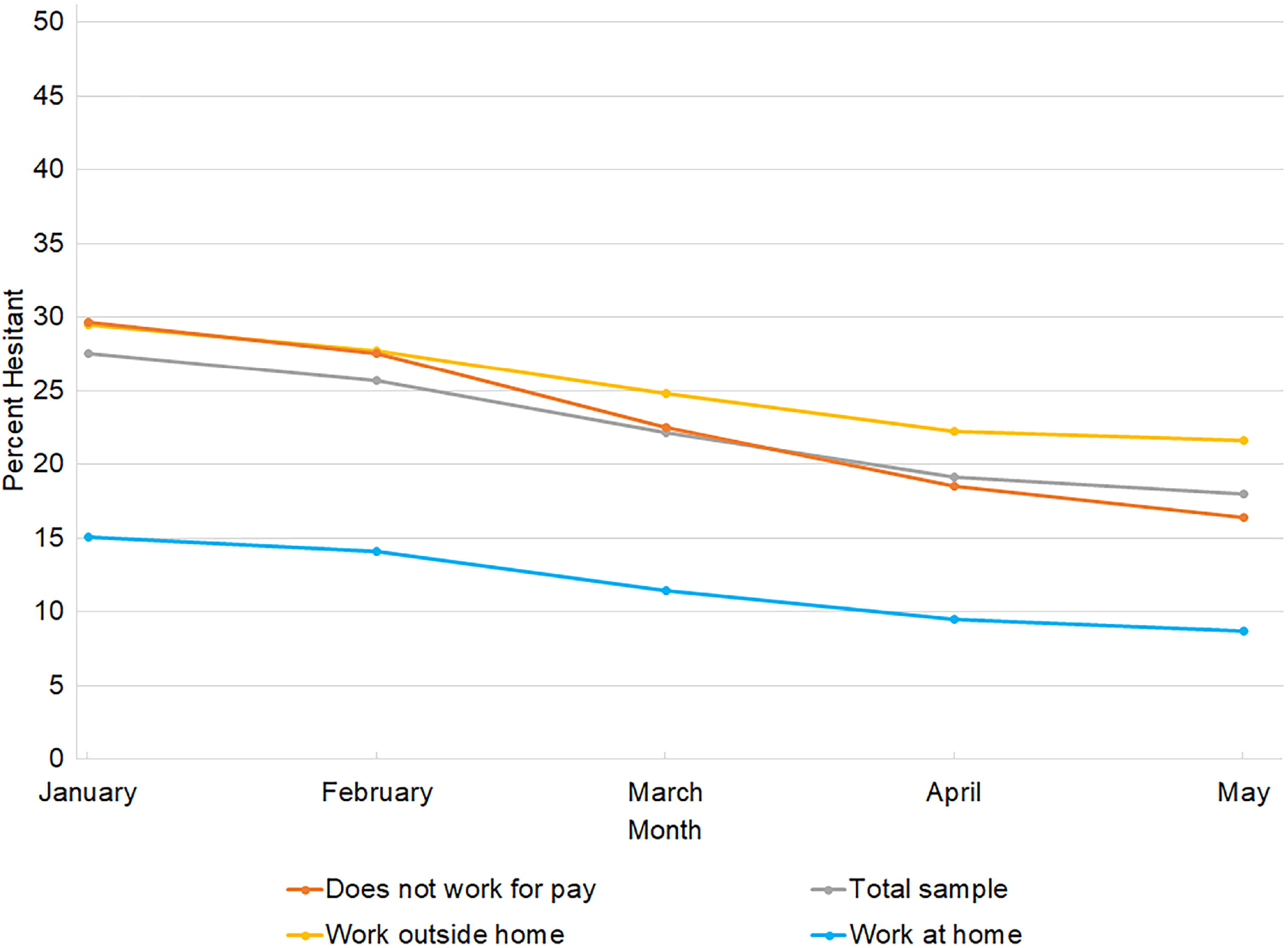
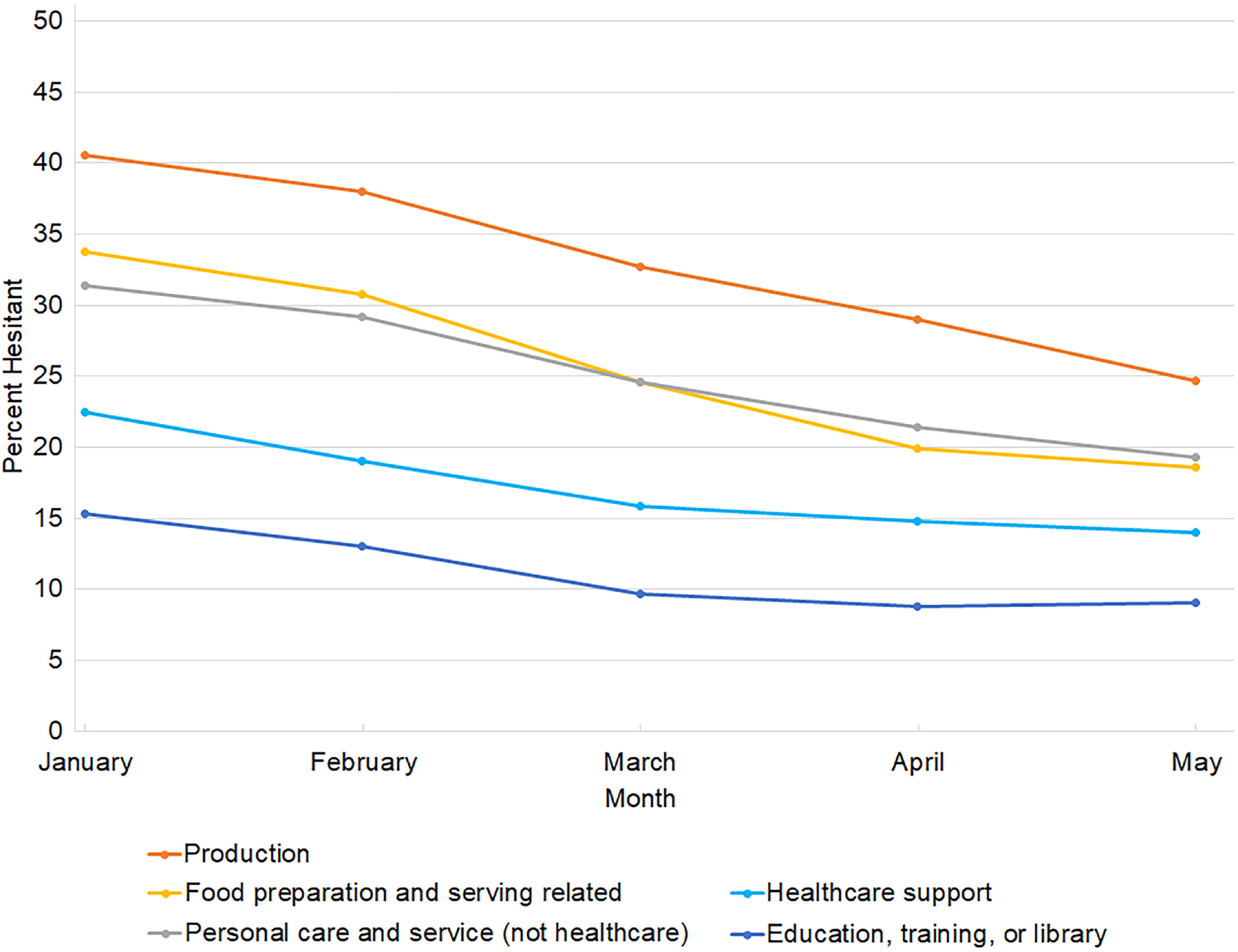
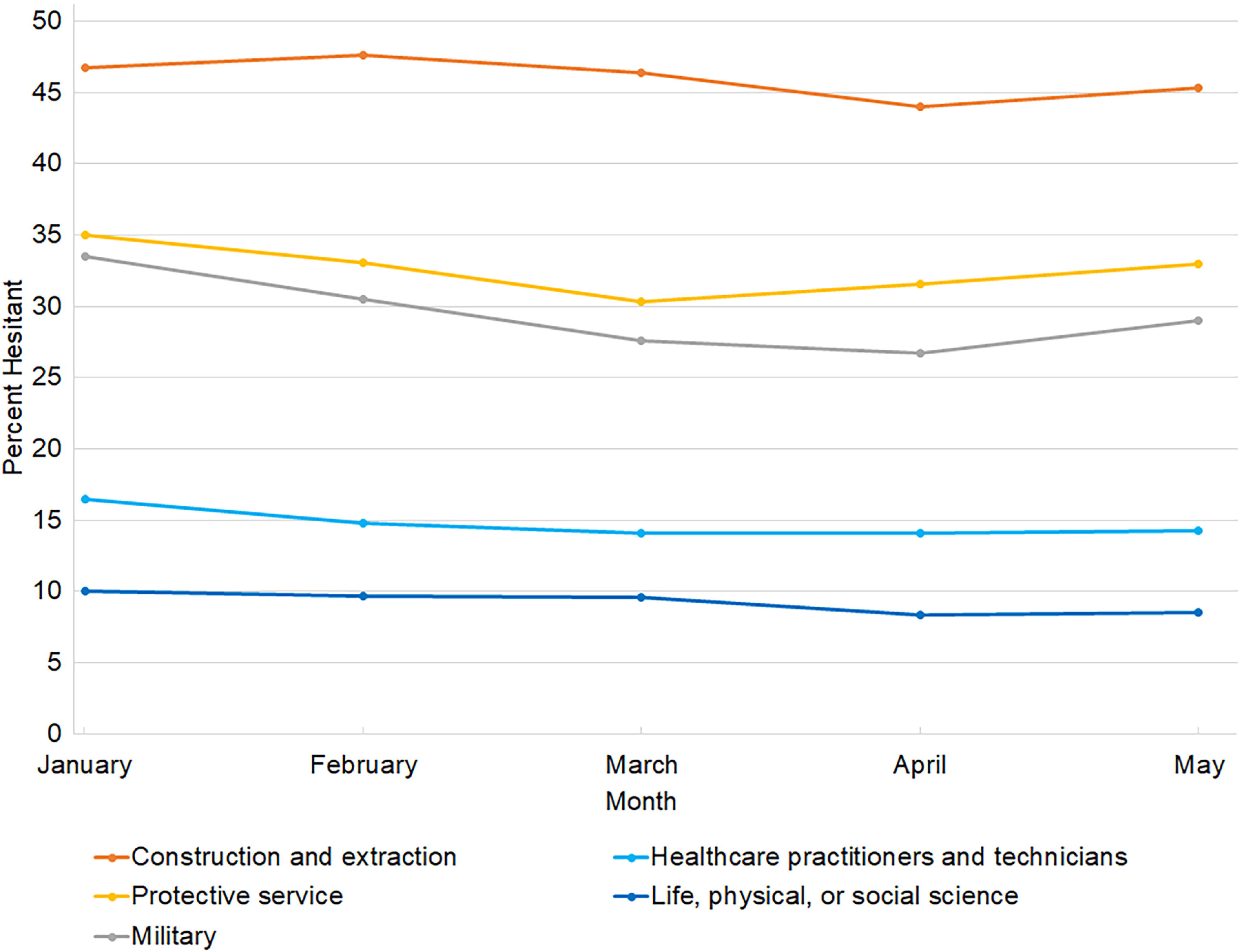
Prevalence of COVID-19 vaccine hesitancy among 18-64 year-old US adults (N=3,179,174) by employment status (A) and select occupational categories with substantial (B) and little change (C), by month, Jan-May 2021. There was a decrease in COVID-19 vaccine hesitancy prevalence between January and May, 2021, among all employment status categories (panel A). There was a smaller change among those who worked outside the home (−7.8%; a 26.6 percent decrease) compared to those who did not work for pay (−13.3%; a 44.9 percent decrease). There was considerable variability in change in prevalence of COVID-19 vaccine hesitancy by occupation category (panels B and C). While most occupations with substantial change in hesitancy had starting points that were relatively high (e.g., production), there were exceptions (e.g., educators) (panel B). Occupations with only small changes in hesitancy from January to May include both those with relatively high starting points (e.g., construction/extraction) and low starting points (e.g., healthcare practitioners/technicians) (panel C).

**Figure 1, panels B and C** shows trajectories of COVID-19 vaccine hesitancy January to May for select occupation categories, highlighting that both the prevalence of hesitancy at a given time point and the degree of change in hesitancy prevalence over time varied by occupation category. While most occupations with substantial change in hesitancy had a relatively high hesitancy prevalence in January (e.g., production, food preparation/serving, personal care/service), there were exceptions (e.g., education/training/library and healthcare support) (**panel B**). Occupations with only small changes in hesitancy from January to May include both those with a relatively high hesitancy prevalence in January (e.g., construction/extraction, protective services, military) and a relatively low prevalence (e.g., healthcare practitioners/technicians, life/physical/social science) (**panel C**). Vaccine hesitancy by month and January to May changes are reported for all occupation categories in **eTable3**.

### April 20 - May 19, 2021

The percentage of respondents who worked outside the home, had received at least one dose of a COVID-19 vaccine, and were COVID-19 vaccine hesitant (strongly and total) during the last 30 days of data collection are reported among employed participants and by occupation category in **Table 1**. Among employed participants, 75.6% [95%CI, 75.5, 75.8]) reported working outside the home. This figure was > 95% in several occupations (e.g., construction/extraction, protective services). However, more than one-third of respondents reported working from home in business/finance operations, management, legal, arts/design/entertainment/sports/media, and office/administrative support. Those in life/physical/social science, and education/training/library occupations led vaccine uptake, which was lowest among those in construction/extraction, installation/maintenance/repair, and farming/fishing/forestry occupations.

**Table 1.** Prevalence of working outside the home, history of COVID-19 vaccination, and COVID-19 vaccine hesitancy,^a^ for employed 18-64 year-old US adults, by occupation category^b^, in May, 2021. Rate ratios for vaccine hesitancy compared to Educators as the reference are also reported.

|  | N | Work outside home<br>% (95% CI) | Vaccinated<br>% (95% CI) | Strongly<br>hesitant<br>% (95% CI) | Hesitant |  |  |
| --- | --- | --- | --- | --- | --- | --- | --- |
|  |  |  |  |  | % (95% CI) | RR (95% CI) | aRR (95% CI) |
| **Employed** | 338226 | 75.6 (75.5, 75.8) | 75.2 (75.0, 75.4) | 13.0 (12.9, 13.2) | 19.0 (18.8, 19.1) | 1.11 (1.09, 1.13) | 1.13 (1.11, 1.15) |
| **Occupation** |  |  |  |  |  |  |  |
| Computer and mathematical | 13047 | 24.7 (23.8, 25.5) | 89.0 (88.3, 89.7) | 4.4 (3.9, 4.8) | 7.3 (6.7, 7.8) | 0.81 (0.74, 0.88) | 0.90 (0.82, 0.98) |
| Life, physical, or social science | 3152 | 66.9 (64.9, 68.9) | 89.1 (87.6, 90.6) | 5.3 (4.3, 6.4) | 8.3 (7.0, 9.5) | 0.92 (0.78, 1.07) | 0.85 (0.72, 0.99) |
| Education, training, or library | 32485 | 78.7 (78.2, 79.3) | 88.7 (88.2, 89.1) | 5.7 (5.4, 6.0) | 9.0 (8.6, 9.3) | Reference | Reference |
| Arts, design, entertainment, sports, and media | 10887 | 53.8 (52.6, 55.0) | 86.3 (85.5, 87.1) | 5.6 (5.1, 6.1) | 9.0 (8.4, 9.7) | 1.01 (0.92, 1.10) | 0.95 (0.87, 1.03) |
| Legal | 4464 | 58.9 (57.2, 60.6) | 87.9 (86.5, 89.2) | 7.2 (6.1, 8.3) | 9.9 (8.6, 11.1) | 1.10 (0.95, 1.25) | 1.07 (0.92, 1.21) |
| Business and finance operations | 10393 | 38.5 (37.3, 39.6) | 83.3 (82.3, 84.2) | 8.0 (7.3, 8.7) | 12.4 (11.5, 13.2) | 1.38 (1.27, 1.49) | 1.42 (1.31, 1.52) |
| Office and administrative support | 44859 | 63.3 (62.8, 63.8) | 83.3 (82.9, 83.8) | 7.9 (7.6, 8.2) | 12.6 (12.2, 13.0) | 1.41 (1.33, 1.48) | 1.32 (1.25, 1.39) |
| Community and social service<sup>c</sup> | 14376 | 80.2 (79.5, 81.0) | 83.1 (82.2, 83.9) | 8.4 (7.8, 9.1) | 13.4 (12.7, 14.2) | 1.50 (1.39, 1.61) | 1.42 (1.32, 1.52) |
| Management | 13319 | 56.6 (55.6, 57.5) | 82.8 (82.0, 83.6) | 10.0 (9.3, 10.7) | 14.3 (13.5, 15.1) | 1.60 (1.49, 1.71) | 1.62 (1.52, 1.73) |
| Healthcare support | 18106 | 78.7 (78.0, 79.4) | 81.5 (80.8, 82.3) | 9.4 (8.8, 9.9) | 14.4 (13.7, 15.0) | 1.61 (1.50, 1.71) | 1.31 (1.23, 1.40) |
| Healthcare practitioners and technicians | 27080 | 93.9 (93.5, 94.2) | 83.3 (82.8, 83.9) | 10.5 (10.0, 10.9) | 14.5 (14.0, 15.0) | 1.62 (1.53, 1.71) | 1.42 (1.34, 1.50) |
| Architecture and engineering | 4559 | 67.0 (65.4, 68.7) | 79.9 (78.3, 81.5) | 11.7 (10.3, 13.0) | 16.5 (15.0, 18.1) | 1.85 (1.66, 2.04) | 1.84 (1.66, 2.01) |
| Food preparation and serving related | 19334 | 96.2 (95.9, 96.5) | 70.3 (69.4, 71.1) | 11.4 (10.8, 12.0) | 19.0 (18.3, 19.8) | 2.13 (2.00, 2.25) | 1.49 (1.41, 1.58) |
| Personal care and service (not healthcare) | 6899 | 82.5 (81.5, 83.6) | 71.9 (70.6, 73.3) | 12.2 (11.3, 13.2) | 19.7 (18.5, 20.8) | 2.20 (2.04, 2.36) | 1.71 (1.59, 1.83) |
| Sales and related | 27095 | 79.3 (78.7, 79.9) | 70.9 (70.2, 71.6) | 14.5 (14.0, 15.1) | 21.8 (21.1, 22.4) | 2.43 (2.31, 2.56) | 1.94 (1.83, 2.04) |
| Building and grounds cleaning/maintenance | 6706 | 94.2 (93.5, 94.9) | 64.7 (63.2, 66.2) | 15.0 (13.9, 16.2) | 22.9 (21.6, 24.2) | 2.56 (2.38, 2.74) | 1.95 (1.81, 2.09) |
| Production<sup>d</sup> | 8409 | 95.5 (95.0, 96.0) | 66.9 (65.7, 68.1) | 17.9 (16.8, 18.9) | 25.9 (24.8, 27.1) | 2.90 (2.71, 3.08) | 2.01 (1.88, 2.13) |
| Military | 1517 | 91.6 (90.1, 93.1) | 66.2 (62.8, 69.6) | 22.3 (19.2, 25.4) | 28.2 (25.0, 31.4) | 3.15 (2.76, 3.53) | 2.15 (1.89, 2.41) |
| Transportation and material moving | 12309 | 95.1 (94.7, 95.6) | 62.1 (61.1, 63.2) | 21.5 (20.6, 22.4) | 29.6 (28.6, 30.6) | 3.31 (3.13, 3.49) | 2.43 (2.29, 2.57) |
| Protective service | 3916 | 95.4 (94.6, 96.3) | 62.7 (60.8, 64.5) | 25.7 (24.0, 27.4) | 32.9 (31.1, 34.6) | 3.67 (3.41, 3.93) | 2.74 (2.55, 2.93) |
| Farming, fishing, and forestry | 2168 | 89.7 (88.4, 91.1) | 53.0 (50.4, 55.7) | 30.7 (28.2, 33.2) | 39.1 (36.5, 41.8) | 4.37 (4.02, 4.72) | 2.66 (2.46, 2.87) |
| Installation, maintenance, repair | 8513 | 95.2 (94.6, 95.7) | 52.1 (50.8, 53.4) | 28.6 (27.4, 29.8) | 39.3 (38.0, 40.6) | 4.39 (4.15, 4.63) | 2.97 (2.81, 3.14) |
| Construction and extraction<sup>e</sup> | 6093 | 96.0 (95.4, 96.6) | 46.6 (45.1, 48.1) | 34.8 (33.2, 36.3) | 45.2 (43.7, 46.8) | 5.05 (4.77, 5.33) | 3.29 (3.10, 3.47) |
| Any other occupation group | 31538 | 78.6 (78.1, 79.1) | 65.1 (64.4, 65.8) | 18.8 (18.2, 19.4) | 26.4 (25.8, 27.0) | 2.95 (2.80, 3.10) | 2.17 (2.06, 2.27) |
| Employed, occupation not reported | 5771 | 0.0 (0.0, 0.0) | 67.2 (65.7, 68.8) | 14.3 (13.2, 15.4) | 21.0 (19.7, 22.2) | 2.34 (2.17, 2.51) | 1.69 (1.54, 1.84) |

<sup>a</sup>Those who reported they would “definitely not” or “probably not” get the vaccine if offered one today were considered vaccine hesitant. Those who reported they would “definitely not” were considered strongly hesitant.
<sup>b</sup>Occupation categories were adapted from the Bureau of Labor Statistics Standard Occupational Classification.
<sup>c</sup>Including counselor, school counselor, mental health worker, social worker, or religious worker.
<sup>d</sup>Including food processing, meat packing, laundry, and dry cleaning workers.
<sup>e</sup>Including oil, gas, mining, or quarrying.

Vaccine hesitancy varied widely by occupation category, with a prevalence of <10% in computer/mathematical, life/physical/social science, education/training/library, and arts/design/entertainment/sports/media, to 25-45% in construction/extraction, installation/maintenance/repair, farming/fishing/forestry, protective services, and transportation/material moving, the military, and production, which includes food processing and meat packing (**Table 1**). Compared to educators, those in construction/extraction had 5 times the chance of vaccine hesitancy (RR=5.05 [95%CI 4.77-5.33]); with adjustment for demographics, including education, they still had >3-fold increased chance (aRR=3.29 [3.10-3.47]).

COVID-19 vaccine hesitancy was similar among health care support (14.4% [95%CI, 14.0-15.0]) and healthcare practitioners (14.5%, [95%CI, 14.0-15.0]). However, hesitancy rates varied by healthcare profession, ranging from 6.9% (95%CI, 4.9-8.8) among pharmacists to 25.2% (95%CI, 21.8-28.6) among emergency medical technicians/paramedics (**Table 2**). Registered nurses and nurse practitioners had relatively low hesitancy (11.6% [10.8-12.3]), while nursing assistant and psychiatric technicians, professions with high patient contact, had a hesitancy prevalence of 18.8% (95%CI, 16.9-20.8); differences in hesitancy between professions were attenuated with control for demographics. Vaccine hesitancy also varied by professions within education/training/library, with a range of 3.6% (95%CI, 2.9-4.2) among post-secondary teachers to 14.8% (13.2-16.3) among preschool/kindergarten teachers.

**Table 2.** Prevalence of working outside the home, history of COVID-19 vaccination, and COVID-19 vaccine hesitancy,^a^ for 18-64 year-old US adults, by profession among health care workers and educators, in May, 2021. Rate ratios for vaccine hesitancy compared to pharmacists and post-secondary teachers, respectively, as the reference are also reported.

|  | N | Work outside<br>home<br>% (95% CI) | Vaccinated<br>% (95% CI) | Strongly<br>hesitant<br>% (95% CI) | Hesitant |  |  |
| --- | --- | --- | --- | --- | --- | --- | --- |
|  |  |  |  |  | % (95% CI) | RR (95% CI) | aRR (95% CI) |
| Healthcare practitioners and support |  |  |  |  |  |  |  |
| Pharmacist | 895 | 92.2 (90.5, 94.0) | 91.9 (89.8, 94.0) | 4.3 (2.7, 6.0) | 6.9 (4.9, 8.9) | Reference | Reference |
| Registered nurse/nurse<br>practioner | 9701 | 93.2 (92.6, 93.7) | 86.7 (85.9, 87.5) | 8.0 (7.4, 8.7) | 11.6 (10.8, 12.3) | 1.68 (1.19, 2.18) | 2.06 (1.45, 2.67) |
| Therapist<sup>b</sup> | 2438 | 94.5 (93.5, 95.5) | 86.6 (85.0, 88.3) | 8.2 (6.9, 9.5) | 12.1 (10.5, 13.7) | 1.76 (1.20, 2.31) | 2.08 (1.42, 2.74) |
| Veterinarian | 647 | 97.6 (96.4, 98.7) | 83.2 (79.6, 86.9) | 9.2 (6.4, 12.1) | 12.8 (9.6, 16.0) | 1.86 (1.15, 2.57) | 1.79 (1.11, 2.47) |
| Physician or surgeon | 1847 | 94.1 (92.9, 95.4) | 85.5 (83.5, 87.6) | 12.6 (10.7, 14.6) | 13.9 (11.9, 15.9) | 2.02 (1.37, 2.67) | 2.28 (1.55, 3.00) |
| Health technologist or<br>technician | 4693 | 94.8 (94.1, 95.5) | 82.9 (81.6, 84.2) | 9.4 (8.4, 10.4) | 14.1 (12.9, 15.3) | 2.05 (1.44, 2.67) | 1.87 (1.30, 2.43) |
| Physician assistant | 656 | 94.6 (92.5, 96.6) | 81.8 (78.2, 85.5) | 12.6 (9.4, 15.8) | 15.8 (12.3, 19.4) | 2.30 (1.47, 3.14) | 2.48 (1.60, 3.37) |
| Dentist | 480 | 94.8 (92.4, 97.1) | 80.9 (76.6, 85.1) | 13.7 (9.8, 17.5) | 17.0 (12.9, 21.1) | 2.47 (1.54, 3.39) | 2.51 (1.61, 3.41) |
| Medical assistant | 1067 | 92.3 (90.3, 94.3) | 78.6 (75.4, 81.8) | 11.6 (9.0, 14.3) | 17.4 (14.5, 20.4) | 2.53 (1.69, 3.38) | 2.22 (1.49, 2.95) |
| Home health or personal<br>care aide | 3349 | 83.7 (82.2, 85.1) | 75.5 (73.6, 77.4) | 11.6 (10.1, 13.2) | 18.0 (16.2, 19.7) | 2.61 (1.82, 3.40) | 2.21 (1.53, 2.88) |
| Licensed practical or<br>licensed vocational nurse | 2180 | 94.7 (93.6, 95.7) | 77.4 (75.3, 79.6) | 13.3 (11.6, 15.0) | 18.9 (16.9, 20.8) | 2.74 (1.90, 3.58) | 2.38 (1.65, 3.11) |
| Nursing assistant or<br>psychiatric aide | 1482 | 95.7 (94.5, 96.9) | 74.4 (71.7, 77.1) | 12.0 (10.1, 13.9) | 19.5 (17.2, 21.9) | 2.84 (1.95, 3.72) | 2.19 (1.50, 2.87) |
| Emergency medical<br>technicians/paramedics | 1073 | 97.0 (95.8, 98.1) | 72.2 (68.7, 75.7) | 20.1 (16.9, 23.3) | 25.3 (21.8, 28.7) | 3.67 (2.50, 4.84) | 2.69 (1.83, 3.54) |
| Other healthcare support | 11568 | 73.9 (73.0, 74.8) | 84.6 (83.8, 85.4) | 8.2 (7.6, 8.9) | 12.4 (11.6, 13.2) | 1.80 (1.27, 2.33) | 1.79 (1.26, 2.32) |
| Other healthcare<br>practitioner | 1654 | 92.9 (91.5, 94.3) | 78.6 (76.2, 81.1) | 14.9 (12.7, 17.2) | 20.0 (17.6, 22.5) | 2.91 (2.00, 3.82) | 3.17 (2.19, 4.16) |
| Educators |  |  |  |  |  |  |  |
| Postsecondary teacher | 4826 | 54.2 (52.5, 55.9) | 94.9 (94.2, 95.7) | 2.5 (1.9, 3.0) | 3.6 (2.9, 4.2) | Reference | Reference |
| Secondary school teacher | 4837 | 91.9 (91.1, 92.8) | 90.3 (89.2, 91.3) | 5.8 (5.0, 6.6) | 8.6 (7.6, 9.5) | 2.40 (1.89, 2.91) | 2.82 (2.22, 3.43) |
| Elementary or middle school teacher | 6712 | 90.2 (89.4, 91.1) | 88.9 (88.0, 89.8) | 6.3 (5.6, 7.0) | 9.6 (8.7, 10.5) | 2.70 (2.16, 3.23) | 3.32 (2.65, 4.00) |
| Preschool or kindergarten teacher | 3112 | 92.6 (91.5, 93.7) | 81.7 (80.0, 83.3) | 9.0 (7.8, 10.3) | 14.8 (13.2, 16.3) | 4.15 (3.29, 5.01) | 3.94 (3.12, 4.77) |
| Other educator<sup>c</sup> | 12002 | 73.9 (72.8, 75.0) | 87.7 (87.0, 88.5) | 5.5 (5.0, 6.0) | 9.0 (8.4, 9.6) | 2.53 (2.04, 3.01) | 2.59 (2.08, 3.09) |

<sup>a</sup>Those who reported they would “definitely not” or “probably not” get the vaccine if offered one today were considered vaccine hesitant. Those who reported they would “definitely not” were considered strongly hesitant.
<sup>b</sup>Including occupational, physical, respiratory, speech
<sup>c</sup>Including teaching assistant, librarian, curator, or other

Reasons for vaccine hesitancy among all employed respondents, and specifically among healthcare workers and educators are reported in **Table 3**. Over half of employed hesitant participants reported concern about side effects (51.7%, 95%CI, 51.1-52.2) and not trusting COVID-19 vaccines (51.3%, 95%CI, 50.8-51.8), whereas only 15.0% (95%CI, 14.6-15.4) didn’t like vaccines in general. Other reasons endorsed by over one-third of respondents were: didn’t believe they needed the vaccine, didn’t trust the government, were waiting to see if the vaccine was safe. The prevalence of reasons for COVID-19 vaccine hesitancy among healthcare practitioners/technicians, healthcare support and educators mostly mirrored that of the overall workforce; however, for all three groups, not trusting the government was a less common reason, while waiting to see if safe, and currently or planning to be pregnant or breastfeeding were more common (**Table 3**).

**Table 3.** Prevalence of reasons for vaccine hesitancy among hesitant^a^ employed 18-64 year-old US adults overall, and for health care practitioners, healthcare support, and educators.

|  | **Total employed** | **HC Practitioners/**<br>**Technicians** | **HC Support** | **Educators** |
| --- | --- | --- | --- | --- |
|  | **N=55375** | **N=3602** | **N=2447** | **N=2580** |
|  | **% (95% CI)** |  |  |  |
| Side effects | 51.7 (51.1, 52.2) | 52.5 (50.5, 54.4) | 54.6 (52.1, 57.1) | 56.3 (54.0, 58.6) |
| Don't trust COVID-19 vaccine | 51.3 (50.8, 51.8) | 48.0 (46.0, 50.0) | 48.9 (46.5, 51.4) | 44.9 (42.7, 47.2) |
| Do not need | 45.1 (44.6, 45.7) | 40.3 (38.3, 42.3) | 36.4 (34.0, 38.8) | 41.7 (39.4, 44.0) |
| Don't trust government | 44.6 (44.1, 45.2) | 35.8 (33.8, 37.7) | 38.3 (35.8, 40.7) | 34.5 (32.3, 36.8) |
| Wait to see if safe then maybe later | 35.2 (34.7, 35.8) | 39.2 (37.2, 41.1) | 42.4 (39.9, 44.9) | 43.8 (41.5, 46.1) |
| Don't know if it will work | 24.2 (23.8, 24.7) | 22.2 (20.5, 23.9) | 24.3 (22.2, 26.4) | 24.3 (22.3, 26.3) |
| Allergic reaction | 22.6 (22.1, 23.0) | 22.0 (20.5, 23.6) | 28.2 (25.9, 30.4) | 24.9 (22.9, 26.9) |
| Don't like vaccines | 15.0 (14.6, 15.4) | 10.8 (9.6, 12.1) | 12.9 (10.8, 15.0) | 13.1 (11.4, 14.8) |
| Other people need more | 14.2 (13.8, 14.6) | 11.9 (10.6, 13.2) | 12.7 (11.2, 14.3) | 14.2 (12.5, 15.8) |
| Doctor not recommended | 9.4 (9.0, 9.7) | 9.3 (8.1, 10.4) | 7.7 (6.5, 8.9) | 10.8 (9.4, 12.2) |
| Safety concern because of health condition | 9.0 (8.7, 9.3) | 11.9 (10.7, 13.2) | 13.3 (11.8, 14.9) | 14.8 (13.1, 16.5) |
| Against religion | 8.6 (8.3, 8.9) | 8.7 (7.6, 9.8) | 8.3 (6.5, 10.1) | 8.6 (7.1, 10.0) |
| Currently/planning to be pregnant/breastfeeding | 6.9 (6.6, 7.1) | 14.3 (12.9, 15.7) | 12.0 (10.3, 13.6) | 13.9 (12.2, 15.5) |
| Cost | 3.3 (3.1, 3.5) | 2.1 (1.5, 2.7) | 2.4 (1.7, 3.1) | 1.9 (1.3, 2.5) |
| Other | 17.1 (16.7, 17.5) | 15.9 (14.5, 17.4) | 12.9 (11.3, 14.5) | 13.4 (11.8, 15.0) |

HC=healthcare
<sup>a</sup>Those who reported they would “definitely not” or “probably not” get the vaccine if offered one today were considered vaccine hesitant.

Reasons for vaccine hesitancy among the five occupations with the highest prevalence of hesitancy are reported in **Table 4**. Compared to all employed hesitant participants, a higher percentage of respondents with jobs in construction/extraction, installation/maintenance/repair, farming/fishing/forestry, protective services, or transportation/material moving reported distrust of the government and not needing the vaccine. With the exception of farming/forestry/fishing, these occupations were also more likely to not trust the vaccine. In contrast, a smaller percentage of those in construction/extraction, and farming/fishing/forestry, reported worry about side effects, an allergic reaction and waiting to see if the vaccine was safe (**Table 4**).

**Table 4.** Prevalence of reasons for COVID-19 vaccine hesitancy among hesitant^a^ 18-64 year-old US adults employed in occupation categories with the highest hesitancy prevalence.

|  | **Construction and**<br>**extraction<sup>a</sup>**<br>**N=2470** | **Installation,**<br>**maintenance, repair**<br>**N=3030** | **Farming, fishing, and**<br>**forestry**<br>**N=770** | **Protective service**<br>**N=1154** | **Transportation and**<br>**material moving<sup>b</sup>**<br>**N=3421** |
| --- | --- | --- | --- | --- | --- |
| Side effects | 43.2 (40.8, 45.7) | 50.4 (48.2, 52.6) | 44.1 (39.7, 48.5) | 52.2 (48.8, 55.6) | 49.6 (47.6, 51.6) |
| Don't trust COVID-19 vaccine | 56.6 (54.2, 59.0) | 55.4 (53.2, 57.6) | 52.9 (48.5, 57.2) | 57.9 (54.6, 61.2) | 56.2 (54.2, 58.2) |
| Do not need | 53.6 (51.1, 56.1) | 56.1 (53.9, 58.3) | 53.0 (48.6, 57.4) | 53.1 (49.7, 56.4) | 49.3 (47.3, 51.3) |
| Don't trust government | 54.8 (52.3, 57.3) | 53.9 (51.7, 56.2) | 52.3 (48.0, 56.7) | 50.7 (47.4, 54.1) | 51.7 (49.7, 53.8) |
| Wait to see if safe then maybe later | 24.0 (21.9, 26.1) | 29.3 (27.3, 31.4) | 23.0 (19.0, 26.9) | 32.8 (29.6, 36.1) | 31.0 (29.1, 32.9) |
| Don't know if it will work | 24.7 (22.5, 26.9) | 24.8 (22.9, 26.6) | 21.4 (17.9, 25.0) | 29.0 (25.9, 32.1) | 25.3 (23.5, 27.1) |
| Allergic reaction | 17.0 (15.2, 18.9) | 19.4 (17.7, 21.2) | 15.1 (12.3, 18.0) | 20.7 (18.1, 23.4) | 21.3 (19.8, 22.9) |
| Don't like vaccines | 15.6 (13.9, 17.3) | 16.9 (15.3, 18.6) | 15.8 (12.5, 19.1) | 13.7 (11.5, 16.0) | 17.3 (15.7, 18.9) |
| Other people need more | 12.1 (10.4, 13.8) | 14.8 (13.2, 16.5) | 10.7 (7.9, 13.5) | 15.0 (12.5, 17.4) | 14.2 (12.6, 15.7) |
| Doctor not recommended | 8.8 (7.4, 10.3) | 10.0 (8.7, 11.3) | 7.6 (5.6, 9.6) | 11.3 (9.1, 13.4) | 9.1 (8.0, 10.3) |
| Safety concern because of health condition | 4.4 (3.5, 5.2) | 5.5 (4.5, 6.4) | 6.1 (4.0, 8.1) | 6.4 (4.9, 7.8) | 6.6 (5.7, 7.4) |
| Against religion | 8.1 (6.9, 9.4) | 8.4 (7.2, 9.6) | 10.4 (7.8, 13.0) | 8.9 (7.0, 10.7) | 10.0 (8.8, 11.2) |
| Currently/planning to be pregnant/breastfeeding | 2.1 (1.4, 2.8) | 1.8 (1.2, 2.4) | 4.6 (2.9, 6.4) | 3.8 (2.5, 5.1) | 2.4 (1.8, 3.1) |
| Cost | 3.5 (2.5, 4.4) | 3.1 (2.3, 3.8) | 2.4 (1.2, 3.6) | 3.5 (2.1, 4.8) | 4.1 (3.2, 4.9) |
| Other | 19.9 (17.9, 21.9) | 18.9 (17.2, 20.5) | 19.5 (16.2, 22.7) | 18.0 (15.5, 20.6) | 17.6 (16.0, 19.2) |

<sup>a</sup>Those who reported they would “definitely not” or “probably not” get the vaccine if offered one today were considered vaccine hesitant.
<sup>a</sup>Including oil, gas, mining, or quarrying
<sup>b</sup>Including delivery services

Reasons for hesitancy among 5 additional occupations selected due to high-density indoor workspace or significant client contact (military, production, food preparation/serving, personal care/service, community and social service) are provided in **eTable4**. Compared to all employed hesitant participants, a higher percentage of respondents in the military reported distrust in the COVID-19 vaccine, disbelief of need and waiting to see if safe; a higher percentage of those in production reported distrust of the government; a higher percentage of those in community and social service or personal care/service reported waiting to see if safe, safety concern because of health conditions and currently/planning to be pregnant or breastfeeding; in addition, among those in personal care/service, concern regarding side effects or an allergic reaction and against religion were more common; finally, a higher percentage of those is food preparation/serving reported concerns regarding side effects or an allergic reaction, waiting to see if safe, other people need more than me, and currently/planning to be pregnant or breastfeeding.

## 4. Discussion

In this massive national survey of adults 18-64 years, COVID-19 vaccine hesitancy decreased by just over one-third from January to May, 2021. While this is a promising finding, 19% of the workforce, and 22% of adults working outside the home in May reported vaccine hesitancy. Furthermore, there was a large disparity in vaccine hesitancy by occupation, with a five-fold difference between the lowest and highest values. While adjustment for demographics reduced the differences in hesitancy between occupation, one-third of occupation categories still had a 2-3.3 fold higher hesitancy that the lowest hesitancy occupations with adjustment. With the emergence of more infectious COVID variants^20^, addressing COVID-19 vaccine hesitancy to improve vaccine uptake is a priority for pandemic control, particularly among the workforce. Occupation categories with the highest hesitancy (construction/extraction, installation/maintenance/repair, farming/fishing/forestry, protective service, and transportation/material moving), include some that have suffered workplace outbreaks, such as agriculture and protective service^8,21^. The majority of hesitant participants in these occupations had strong hesitancy (i.e., responded “definitely not”) and reported not trusting the government and/or the COVID-19 vaccine, indicating that their hesitancy may be based in strong beliefs about the government or the vaccine development process. Further, they were more likely than all employed hesitant participants to believe they do not need the vaccine. In some of these professions, individuals may work primarily outside or in uncrowded conditions and feel less at risk of contracting COVID-19. Thus, their reasons for hesitancy indicate a need for public health campaigns to increase trust in the COVID-19 vaccine and the government, and to increase awareness of the benefits of a COVID-19 vaccine to employees and their community in order to address the belief that some individuals do not need the vaccine.

Given the variation in hesitancy by occupational groups, public health and medical workers could seek to understand and address reasons for hesitancy in specific workplace communities by building partnerships in occupations with high vaccine hesitancy. Workplace vaccination clinics have the potential to address several potential barriers to COVID-19 vaccination, e.g., difficulty scheduling, transportation, travel and time requirements, including unpaid time off of work, and of going to an unfamiliar location^22–24^. In addition to clinics, employers can promote vaccine access by ensuring paid time off and offering transportation to workers to receive vaccines offsite. Workplace efforts can address poor understanding of the risks and benefits, and lack of vaccination being the norm, by providing population-specific educational messaging and positive peer pressure^22–24^. The Centers for Disease Control and Prevention (CDC) provides COVID-19 vaccination audience-specific toolkits to promote vaccine acceptance, including an essential workers toolkit^25^, and guidance to employers on hosting workplace vaccination clinics^26^. They advise including management, human resources, employees and labor representatives, as appropriate, in the planning process, and using multiple strategies to promote and encourage participation in the vaccination clinics, e.g., encouraging managers and leaders to get vaccinated first. Just as celebrities have promoted vaccination to the public and Black health care workers have had success engaging Black communities^27^, workplace-focused campaigns could feature prominent and ordinary figures from specific workplaces or occupations discussing why they got vaccinated^28^.

Among healthcare workers, several professions with high patient contact (e.g., nursing assistants/psychiatric aides) reported hesitancy >15%. This is concerning as patients are often at higher risk of hospitalization or death from COVID-19 than the general population, based on their age or health status. Published guidance on promoting vaccinations among healthcare workers^29,30^ may serve as a starting point for COVID-19 specific efforts.

Hesitancy among educators was generally low. However, 15% of preschool and kindergarten teachers and 10% of elementary school teachers, whom teach children not yet eligible for a COVID-19 vaccine were hesitant. While some universities and private schools are requiring students, staff and faculty to be vaccinated before the start of the 2021 fall semester^31,32^, most private and public preschool and elementary schools have no vaccine mandates^33^. Many also lack masking mandates^34^, making vaccination even more important.

A striking finding was that participants working outside the home reported COVID-19 vaccine hesitancy at more than twice the rate of those working from home. This may reflect the observed difference in hesitancy by occupation, as working from home was more common in some occupations than others. This finding may also reflect that those who are more worried about COVID-19, who as a group have less COVID-19 vaccine hesitancy^35^, are choosing to work from home when possible.

### Study limitations and strengths

Cross-sectional samples were used to evaluate time trends, and the sample representativeness may have been affected by the recruitment method and low response rate. Specifically, this study used a novel sampling method with a soft ask. Responses were weighted to match the age, gender, and state profile of the US population^7^, but representativeness within each occupational category is not guaranteed. Additionally, studies from the previous decade found differences in personality traits between Facebook users and non-users^36,37^. While we do not expect those exact findings to hold a decade later in the much larger and more diverse Facebook user population^38^, the Facebook user and general US populations are expected to differ, and we could not control for unmeasured differences between them or the impact of receiving vaccine-related content through Facebook itself. Compared to the American Community Survey 2015-2019 5-year 2 Data Release^39^, demographics of the weighted sample are similar to the US population, but white race and higher education are slightly over-represented, and vaccine uptake is over-represented^3^. Thus, overall hesitancy prevalence estimates were likely underestimated. A study strength is that vaccinated individuals were included in the vaccine accepting (i.e., not hesitant) group, as access to vaccination varied by occupational group over the time studied. Thus, assessment of time trends or comparisons between occupation categories should be valid.

Additional study strengths include the timing of our study (i.e., during the first five months of the COVID-19 vaccine rollout) and our large geographically and occupationally diverse sample, which allowed for comparisons by month and occupation. This large-scale national sample with detailed data on occupational categories and respondent characteristics is, to the author’s knowledge, the best US data available on COVID-19 hesitancy by employment and occupation.

## 5. Conclusions

Vaccine hesitancy among US adults 18-64 years decreased in the first five months of the US COVID-19 vaccine rollout. However, with approximately one in five members of the US workforce hesitant in May, 2021, and some occupational categories reporting hesitancy at twice this rate, vaccine hesitancy remains a threat to COVID-19 pandemic control. This report identified occupations with high rates of COVID-19 vaccine hesitancy and awthe workforce and in specific occupations to help public health and health care workers target interventions and address specific concerns to increase vaccination rates, potentially via workplace-focused campaigns and onsite vaccination clinics. Messaging about safety, addressing trust, and clarifying the value of vaccinations to prevent COVID-19 is needed.

## Data Availability

If you are interested in using the survey data for your research, you can start the process by submitting a form requesting a data use agreement (DUA) from Facebook. The data is not available from the authors.

https://dataforgood.fb.com/docs/covid-19-symptom-survey-request-for-data-access/

## Acknowledgements

The authors would like to thank the Delphi Group at Carnegie Mellon University for input and support on the survey instrument. Wichada La Motte-Kerr, MPH, of Delphi contributed to the development and deployment of the survey and received compensation for her contributions to the study. We thank Sarah LaRocca, PhD and Katherine Morris, PhD of Facebook for contributions to the survey instrument design.

