## Supplemental Material for "COVID-19 vaccine hesitancy January-May 2021 among 18-64 year old US adults by employment and occupation"

**Appendix.** The COVID-19 Trends and Impact Survey (CTIS): select questions and response sets.

The survey questions used in this report are provided below. A listing of professions by occupation category, which were adapted from the Bureau of Labor Statistics Standard Occupational Classification, is provided following the survey questions. The full questionnaire and revisions history can be seen online: https://cmu-delphi.github.io/delphi-epidata/symptom-survey/coding.html

D1 What is your gender?

1. Male
2. Female
3. Non-binary
4. Prefer to self-describe:
5. Prefer not to answer

D2 What is your age?

1. 18-24 years
2. 25-34 years
3. 35-44 years
4. 45-54 years
5. 55-64 years
6. 65-74 years
7. 75 years or older

D6 Are you of Hispanic, Latino, or Spanish origin?

1. Yes
2. No, not of Hispanic, Latino, or Spanish origin

D7 What is your race? Please select all that apply.

1. American Indian or Alaska Native
2. Asian
3. Black or African American
4. Native Hawaiian or other Pacific Islander
5. White
6. Some other race

Continued next page

D8 What is the highest degree or level of school you have completed?

1. Less than high school
2. High school graduate or equivalent (GED)
3. Some college
4. 2 year degree
5. 4 year degree
6. Master's degree
7. Professional degree (e.g. MD, JD, DVM)
8. Doctorate

D9 In the past 4 weeks, did you do any kind of work for pay?

1. Yes
2. No

D10 [Displayed if D9 = “Yes”] Was any of your work for pay in the last four weeks outside your home?

1. Yes
2. No

Q64 [Displayed if D9 = “Yes”] Please select the occupational group that best fits the main kind of work you were doing in the last four weeks.

1. Community and social service (including counselor, school counselor, mental health worker, social worker, or religious worker)
2. Education, training, and library
3. Arts, design, entertainment, sports, and media
4. Healthcare practitioners and technicians
5. Healthcare support
6. Protective service
7. Food preparation and serving related (including grocery store workers)
8. Building and grounds cleaning and maintenance
9. Personal care and service (not healthcare)
10. Sales and related
11. Office and administrative support (including postal workers)
12. Construction and extraction (oil, gas, mining, or quarrying)
13. Installation, maintenance, and repair
14. Production (including food processing, meat packing, laundry, and dry cleaning workers)
15. Transportation and material moving (including delivery services)
16. Other occupation

Q66 [Displayed if Q64 = "2. Education, training, and library"] Please select the job type that best fits the main kind of work you were doing in the last four weeks.

1. Preschool or kindergarten teacher
2. Elementary or middle school teacher
3. Secondary school teacher
4. Postsecondary teacher
5. Other teacher or instructor, including special education
6. Teacher assistant
7. Librarian, library technician, archivist, curator, or museum technician

Q68 [Displayed if Q64 = "4. Healthcare practitioners and technicians"] Please select the job type that best fits the main kind of work you were doing in the last four weeks.

1. Physician or surgeon
2. Registered nurse (including nurse practitioner)
3. Licensed practical or licensed vocational nurse
4. Physician assistant
5. Dentist
6. Any other treating practitioner (chiropractor, dietitian or nutritionist, optometrist, podiatrist, audiologist, acupuncturist, dental hygienist)
7. Pharmacist
8. Any therapist (occupational, physical, respiratory, speech)
9. Any health technologist or technician (including hospital laboratory scientist and pharmacy technician)
10. Veterinarian
11. Emergency medical technicians and paramedics

Q69 [Displayed if Q64 = "5. Healthcare support"] Please select the job type that best fits the main kind of work you were doing in the last four weeks.

1. Nursing assistant or psychiatric aide
2. Home health or personal care aide (including in-home caregivers)
3. Occupational therapy or physical therapist assistant or aide
4. Massage therapist
5. Dental assistant
6. Medical assistant
7. Medical transcriptionist
8. Pharmacy aide
9. Phlebotomist
10. Veterinary assistant or laboratory animal caretaker
11. Any other healthcare support worker, including medical equipment preparer

Continued on the next page

Q80 [Displayed if Q64 = “16. Other occupation”] Please select the occupational group that best fits the main kind of work you were doing in the last four weeks.

Other occupation

1. Management
2. Business and financial operations
3. Computer and mathematical
4. Architecture and engineering
5. Life, physical, and social science
6. Legal
7. Farming, fishing, and forestry
8. Military
9. Any other occupational group

V1 Have you had a COVID-19 vaccination?

1. Yes
2. No
3. I don't know

[Displayed if V1 is not “Yes”] If a vaccine to prevent COVID-19 were offered to you today, would you choose to get vaccinated?

1. Yes, definitely
2. Yes, probably
3. No, probably not
4. No, definitely not

V5a-c :

V5a [Displayed if V3 = “Yes, probably”] Which of the following, if any, are reasons that you only probably will* get a COVID-19 vaccine? Please select all that apply.

V5b [Displayed if V3 = “No, probably not”] Which of the following, if any, are reasons that you probably won't* get a COVID-19 vaccine? Please select all that apply.

V5c [Displayed if V3 = “No, definitely not”] Which of the following, if any, are reasons that you definitely won't* get a COVID-19 vaccine? Please select all that apply.

*The wording of the response sets was changed from “will get” to “would choose to get,” and “won’t get” to “wouldn’t choose to get,” on February 8, 2021.

Continued on the next page

Answer choices for both versions:

1. I am concerned about possible side effects of a COVID-19 vaccine.
2. I am concerned about having an allergic reaction to a COVID-19 vaccine.
3. I don't know if a COVID-19 vaccine will work.
4. I don't believe I need a COVID-19 vaccine.
5. I don't like vaccines.
6. My doctor has not recommended it.
7. I plan to wait and see if it is safe and may get it later.
8. I think other people need it more than I do right now.
9. I am concerned about the cost of a COVID-19 vaccine.
10. I don't trust COVID-19 vaccines.
11. I don't trust the government.
12. It is against my religious beliefs.
13. I have a health condition and am concerned about the safety of the vaccine for people with my condition.
14. I am currently/planning to be pregnant and/or breastfeeding and do not want to get vaccinated at this time.
15. Other

**Examples of professions by occupation category available to survey respondents were:**

Arts, design, entertainment, sports, and media

- Art worker (fine, craft, multimedia)
- Design worker (fashion, floral, graphic, interior, set and exhibit)
- Entertainer or performer (actor, producer, director, dancer, choreographer, singer, musician)
- Sports and related worker (athlete, coach, scout, umpire, referee)
- Media and communication worker (announcer, analyst, report, editor, translator)
- Media and communication equipment worker (audio or video technician)
- Any other arts, design, entertainment, sports, or media worker

Building and grounds cleaning and maintenance

- First-line supervisor of housekeeping or janitorial workers
- First-line supervisor of landscaping, lawn service, or groundskeeping workers
- Janitor or building cleaner
- Maid or housekeeping cleaner
- Pest control worker
- Grounds maintenance worker
- Any other building and grounds cleaning or maintenance worker

Community and social service

- Counselor
- Social worker
- Social or human service assistant
- Probation officer or correctional treatment specialist
- Clergy or other religious worker
- Any other community or social service specialist

Construction and extraction

- First-line supervisor of construction trades or extraction workers
- Any construction trades worker (carpenter, electrician, plumber, roofer, helper)
- Any other construction worker, including inspector and highway worker
- Any extraction worker in oil, gas, mining, or quarrying

Education, training, and library

- see Q66

Food preparation and serving related

- Chef, head cook, or first-line supervisor of food preparation and serving workers
- Cook
  Food preparation worker  Bartender  Fast food or counter worker
- Waiter or waitress
- Food server, non-restaurant
  Dining room or cafeteria attendant or bartender helper  Dishwasher
- Host or hostess at a restaurant, lounge, or coffee shop
- Any other food preparation and serving related worker
- Grocery store worker

Healthcare practitioners and technicians

- see Q68

Healthcare support

- see Q69

Continued on the next page

Installation, maintenance, and repair

- First-line supervisor of mechanics, installers, or repairers
- Electrical or electronic equipment mechanic, installer, or repairer
- Vehicle or mobile equipment mechanic, installer, or repairer (aircraft, automotive, bus, truck, heavy vehicles)
- Heating, air conditioning, and refrigeration mechanic or installer
- Line installer or repairer (electrical or telecommunications)
- Any other installation, maintenance, or repair worker

Office and administrative support

- First-line supervisor of office or administrative support workers
- Financial clerk including bookkeeping, accounting, auditing, or billing
- Customer service representative
- Receptionist or information clerk
- Postal service worker or mail carrier
- Shipping, receiving, or inventory clerk
- Secretary or administrative assistant
- Any other office or administrative support worker

Personal care and service (not healthcare)

- Hairdresser, hairstylist, cosmetologist, or barber
- Any other personal appearance worker
- Childcare worker
- Animal care or training worker
- Gambling service worker
- Miscellaneous entertainment attendant
- Funeral service worker
- Recreation or fitness worker
- Any other personal care or service worker

Continued on the next page

Production

- First-line supervisor of production and operating workers
- Any assembler or fabricator
- Food processing worker
- Metal or plastic worker (machinist, welder, soldering)
- Printing worker
- Laundry or dry-cleaning worker
- Any other textile, apparel, or furnishings worker
- Woodworker
- Plant and system operator (power, water, wastewater, chemical)
- Any other production worker

Protective services

- First-line supervisor (firefighter, police, correctional, or security)
- Firefighter, fire inspector, or fire investigator
- Police or sheriff officer
- Detective or criminal investigator
- Bailiff, correctional officer, or jailer
- Security guard or gaming surveillance officer
- Lifeguard, ski patrol, or other recreational protective service worker
- Any other protective service worker

Sales and related

- First-line supervisor of sales workers
- Cashier
- Retail salesperson (including counter or rental clerk or parts salesperson)
- Sales representative in services, wholesale, or manufacturing
- Real estate broker or sales agent
- Telemarketer
- Any other sales or related worker

Transportation and material moving

- First-line supervisor of transportation or material moving workers
- Air transportation worker (pilot, flight engineer, air traffic controller, flight attendant)
- Motor vehicle operator
- Rail transportation worker (including railway, subway, and streetcar operator)
- Water transportation worker
- Any other transportation worker
- Any material moving worker

No examples were included in the survey instrument for the following occupations:

- Management
- Business and financial operations
- Computer and mathematical
- Architecture and engineering
- Life, physical, and social science
- Legal
- Farming, fishing, and forestry
- Military

**eTable 1.** Respondent flow for study of COVID-19 vaccine hesitancy in 18-64 year old US adults, by month of survey completion[a]

| | January | February | March | April | May | Total |
|---|---|---|---|---|---|---|
| | N | | | | | |
| Offered Survey | 106387320 | 95953902 | 104768154 | 103399752 | 104760491 | 515269619 |
| Responded (≥2 items) | 1314126 | 1232691 | 1291957 | 1084516 | 538910 | 5462200 |
| Age 65 and over | 287811 | 302590 | 334570 | 268657 | 132883 | 1326511 |
| Did not report age or hesitancy | 234599 | 219572 | 225079 | 184238 | 93027 | 956515 |
| Report sample | 791716 | 710529 | 732308 | 631621 | 313000 | 3179174 |
| Did not report employment | 9325 | 7649 | 7175 | 6571 | 3457 | 34177 |

[a]Data was collected continuously from January 6 to May 19, 2021, and aggregated by month to evaluate time trends.

**eTable 2**. Past month employment status and COVID-19 vaccine hesitancy overall and by employment status among 18-64 year-old US adults, by month, Jan-May 2021

| | January N=791716 | February N=710529 | March N=732308 | April N=631621 | May N=313000 | Difference May - Jan | Percent change (May – Jan)/Jan |
|---|---|---|---|---|---|---|---|
| | **% (95% CI)** | | | | | | |
| **Employment status** | | | | | | | |
| No response | 2.5 (2.4, 2.5) | 2.3 (2.3, 2.3) | 2.2 (2.1, 2.2) | 2.3 (2.2, 2.3) | 2.3 (2.2, 2.4) | -0.2 (-0.2, -0.1) | -6.6 (-9.8, -3.4) |
| Did not work for pay | 33.7 (33.6, 33.8) | 33.9 (33.8, 34.0) | 33.2 (33.1, 33.3) | 33.2 (33.1, 33.4) | 33.3 (33.1, 33.5) | -0.4 (-0.6, -0.1) | -1.0 (-1.8, -0.3) |
| Worked for pay | 65.0 (64.9, 65.1) | 64.9 (64.7, 65.0) | 65.7 (65.6, 65.8) | 65.6 (65.5, 65.7) | 65.5 (65.3, 65.7) | 0.5 (0.3, 0.7) | 0.8 (0.4, 1.1) |
| Worked outside home | 48.1 (48.0, 48.2) | 48.7 (48.6, 48.9) | 49.7 (49.5, 49.8) | 49.4 (49.2, 49.5) | 49.8 (49.5, 50.0) | 1.7 (1.4, 1.9) | 3.5 (3.0, 4.0) |
| Worked at home | 15.8 (15.7, 15.9) | 15.1 (15.0, 15.2) | 14.9 (14.9, 15.0) | 15.1 (15.0, 15.2) | 14.6 (14.5, 14.7) | -1.2 (-1.3, -1.0) | -7.4 (-8.5, -6.3) |
| **COVID-19 vaccine hesitancy** | | | | | | | |
| Total sample | 27.5 (27.3, 27.6) | 25.7 (25.6, 25.8) | 22.1 (21.9, 22.2) | 19.1 (19.0, 19.2) | 18.0 (17.8, 18.2) | -9.5 (-9.7, -9.3) | -34.5 (-35.2, -33.8) |
| Employment status | | | | | | | |
| No response | 36.8 (35.9, 37.7) | 33.3 (32.3, 34.2) | 27.3 (26.4, 28.3) | 23.5 (22.5, 24.4) | 22.0 (20.8, 23.3) | -14.8 (-16.3, -13.2) | -40.1 (-43.9, -36.3) |
| Did not work for pay | 29.7 (29.5, 29.9) | 27.5 (27.3, 27.7) | 22.5 (22.3, 22.7) | 18.5 (18.3, 18.7) | 16.4 (16.1, 16.6) | -13.3 (-13.7, -13.0) | -44.9 (-45.9, -43.9) |
| Worked for pay | 26.1 (26.0, 26.3) | 24.6 (24.5, 24.8) | 21.8 (21.6, 21.9) | 19.3 (19.2, 19.5) | 18.8 (18.5, 19.0) | -7.4 (-7.6, -7.1) | -28.2 (-29.1, -27.2) |
| Work outside home | 29.5 (29.3, 29.7) | 27.7 (27.5, 27.9) | 24.8 (24.6, 24.9) | 22.2 (22.0, 22.4) | 21.6 (21.4, 21.9) | -7.8 (-8.2, -7.5) | -26.6 (-27.7, -25.6) |
| Work at home | 15.1 (14.9, 15.3) | 14.1 (13.8, 14.3) | 11.4 (11.2, 11.6) | 9.5 ( 9.3, 9.7) | 8.7 ( 8.4, 9.0) | -6.4 (-6.8, -6.1) | -42.6 (-44.9, -40.3) |

Jan=January

**eTable 3.** COVID-19 vaccine hesitancy in 18-64 year-old US adults by occupation category, by month January-May 2021.

| | January | February | March | April | May | Difference May - Jan | Percent change (May – Jan)/Jan |
|---|---|---|---|---|---|---|---|
| **Occupation category** | % (95%CI) | | | | | | |
| Architecture and engineering | 16.8 (15.6, 18.0) | 20.8 (19.3, 22.2) | 18.3 (17.1, 19.5) | 16.4 (15.1, 17.8) | 16.0 (14.1, 17.9) | -0.8 (-3.0, 1.5) | -4.7 (-17.9, 8.5) |
| Arts, design, entertainment, sports, and media | 13.5 (12.8, 14.1) | 13.7 (12.9, 14.4) | 12.8 (12.1, 13.6) | 9.9 (9.3, 10.6) | 8.9 (8.1, 9.8) | -4.5 (-5.6, -3.4) | -33.6 (-40.7, -26.4) |
| Building and grounds cleaning and maintenance | 32.2 (31.1, 33.4) | 32.4 (31.2, 33.7) | 28.4 (27.1, 29.6) | 24.3 (23.0, 25.6) | 22.7 (21.1, 24.3) | -9.6 (-11.5, -7.6) | -29.7 (-35.2, -24.1) |
| Business and finance operations | 17.4 (16.7, 18.2) | 16.9 (16.2, 17.7) | 15.3 (14.6, 16.1) | 12.9 (12.1, 13.8) | 12.4 (11.3, 13.5) | -5.0 (-6.3, -3.7) | -28.8 (-35.7, -21.9) |
| Community and social service[a] | 19.5 (18.8, 20.1) | 16.5 (15.9, 17.1) | 14.0 (13.4, 14.6) | 13.0 (12.4, 13.7) | 13.4 (12.4, 14.4) | -6.1 (-7.2, -4.9) | -31.1 (-36.7, -25.5) |
| Computer and mathematical | 12.1 (11.5, 12.6) | 12.0 (11.3, 12.6) | 10.3 (9.7, 10.9) | 7.7 (7.2, 8.2) | 7.5 (6.7, 8.2) | -4.6 (-5.6, -3.6) | -38.0 (-45.2, -30.9) |
| Construction and extraction | 46.7 (45.4, 48.0) | 47.6 (46.2, 48.9) | 46.4 (45.1, 47.7) | 44.0 (42.6, 45.4) | 45.3 (43.2, 47.3) | -1.4 (-3.8, 1.0) | -3.0 (-8.1, 2.0) |
| Education, training, or library | 15.3 (14.9, 15.6) | 13.0 (12.6, 13.3) | 9.7 (9.4, 10.1) | 8.8 (8.5, 9.1) | 9.1 (8.5, 9.6) | -6.2 (-6.8, -5.6) | -40.7 (-44.3, -37.1) |
| Farming, fishing, and forestry | 43.4 (41.0, 45.8) | 44.1 (41.6, 46.6) | 41.2 (38.9, 43.5) | 38.4 (36.0, 40.8) | 38.7 (35.4, 42.0) | -4.7 (-8.8, -0.7) | -10.9 (-19.9, -1.8) |
| Food preparation and serving related | 33.8 (33.1, 34.5) | 30.8 (30.1, 31.5) | 24.6 (23.9, 25.2) | 19.9 (19.2, 20.5) | 18.6 (17.6, 19.6) | -15.2 (-16.5, -14.0) | -45.1 (-48.3, -41.9) |
| Healthcare practitioners and technicians | 16.5 (16.1, 16.9) | 14.8 (14.5, 15.2) | 14.1 (13.7, 14.5) | 14.1 (13.6, 14.6) | 14.3 (13.6, 15.0) | -2.2 (-3.0, -1.4) | -13.2 (-17.8, -8.6) |
| Healthcare support | 22.5 (22.0, 23.0) | 19.0 (18.5, 19.5) | 15.9 (15.4, 16.4) | 14.8 (14.3, 15.4) | 14.0 (13.2, 14.9) | -8.5 (-9.5, -7.5) | -37.6 (-41.7, -33.5) |
| Installation, maintenance, repair | 41.8 (40.8, 42.9) | 43.7 (42.6, 44.9) | 42.6 (41.5, 43.7) | 40.3 (39.0, 41.5) | 38.3 (36.7, 40.0) | -3.5 (-5.4, -1.5) | -8.3 (-12.9, -3.8) |
| Legal | 12.0 (11.0, 12.9) | 11.5 (10.5, 12.6) | 10.0 (9.1, 11.0) | 9.6 (8.5, 10.7) | 9.6 (8.0, 11.2) | -2.4 (-4.2, -0.5) | -19.9 (-34.4, -5.3) |

| | | | | | | | |
|---|---|---|---|---|---|---|---|
| Life, physical, or social science | 10.0 (8.9, 11.1) | 9.7 (8.6, 10.8) | 9.6 (8.5, 10.7) | 8.4 (7.2, 9.6) | 8.5 (6.7, 10.2) | -1.6 (-3.6, 0.5) | -15.7 (-35.2, 3.9) |
| Management | 18.7 (18.0, 19.3) | 19.0 (18.3, 19.7) | 17.5 (16.8, 18.2) | 13.9 (13.2, 14.6) | 14.6 (13.7, 15.6) | -4.0 (-5.2, -2.8) | -21.5 (-27.6, -15.5) |
| Military | 33.5 (31.1, 35.9) | 30.5 (28.0, 33.0) | 27.6 (25.2, 30.1) | 26.7 (23.9, 29.5) | 29.0 (24.8, 33.2) | -4.5 (-9.3, 0.3) | -13.4 (-27.4, 0.5) |
| Office and administrative support | 20.7 (20.4, 21.1) | 19.3 (18.9, 19.7) | 16.1 (15.7, 16.4) | 12.9 (12.6, 13.3) | 12.3 (11.8, 12.8) | -8.5 (-9.1, -7.8) | -40.8 (-43.4, -38.2) |
| Personal care and service (not healthcare) | 31.4 (30.4, 32.4) | 29.2 (28.2, 30.3) | 24.6 (23.6, 25.7) | 21.4 (20.3, 22.5) | 19.3 (17.9, 20.8) | -12.0 (-13.8, -10.3) | -38.4 (-43.4, -33.3) |
| Production[b] | 40.6 (39.7, 41.5) | 38.0 (37.0, 39.1) | 32.7 (31.6, 33.8) | 29.0 (27.9, 30.2) | 24.7 (23.2, 26.1) | -15.9 (-17.7, -14.2) | -39.3 (-43.1, -35.4) |
| Protective service | 35.0 (33.6, 36.4) | 33.1 (31.6, 34.5) | 30.3 (28.9, 31.7) | 31.6 (29.9, 33.2) | 33.0 (30.7, 35.3) | -2.0 (-4.7, 0.7) | -5.6 (-13.2, 2.0) |
| Sales and related | 30.0 (29.4, 30.5) | 29.5 (28.9, 30.1) | 26.9 (26.3, 27.4) | 23.1 (22.5, 23.7) | 21.3 (20.5, 22.2) | -8.6 (-9.6, -7.6) | -28.8 (-31.8, -25.8) |
| Transportation and material moving | 38.5 (37.7, 39.4) | 37.1 (36.2, 38.0) | 34.0 (33.1, 34.9) | 31.3 (30.4, 32.3) | 29.7 (28.5, 31.0) | -8.8 (-10.3, -7.3) | -22.8 (-26.6, -19.1) |
| Other occupation group | 34.4 (33.8, 34.9) | 33.1 (32.6, 33.7) | 30.9 (30.3, 31.4) | 27.1 (26.5, 27.7) | 25.8 (25.0, 26.6) | -8.6 (-9.6, -7.7) | -25.1 (-27.7, -22.5) |
| Employed, occupation not reported | 36.9 (35.7, 38.1) | 33.3 (32.0, 34.6) | 27.7 (26.5, 29.0) | 23.0 (21.7, 24.2) | 21.3 (19.7, 22.9) | -15.5 (-17.6, -13.5) | -42.2 (-47.0, -37.4) |

Jan=January

[a]Including counselor, school counselor, mental health worker, social worker, or religious worker.

[b]Including food processing, meat packing, laundry, and dry cleaning workers.

**eTable 4.** Reasons for vaccine hesitancy among hesitant 18-64 year-old US adults in occupation categories with high-density indoor workspaces or significant client contact

| | Military N=360 | Production[a] N=1995 | Personal care and service[b] N=1322 | Food prep/serving N=3669 | Community and social service N=1650 |
|---|---|---|---|---|---|
| **Reasons** | **% (95% CI)** | | | | |
| Side effects | 55.3 (48.4, 62.1) | 52.7 (50.0, 55.3) | 57.6 (54.4, 60.7) | 55.9 (53.7, 58.1) | 53.2 (50.1, 56.2) |
| Don't trust COVID-19 vaccine | 60.1 (53.5, 66.7) | 54.2 (51.6, 56.9) | 48.8 (45.5, 52.0) | 50.6 (48.4, 52.9) | 44.3 (41.2, 47.4) |
| Do not need | 57.8 (51.0, 64.5) | 48.1 (45.4, 50.8) | 35.2 (32.1, 38.2) | 37.3 (35.0, 39.5) | 40.5 (37.5, 43.6) |
| Don't trust government | 45.1 (38.1, 52.1) | 49.2 (46.5, 51.9) | 40.1 (36.9, 43.3) | 43.0 (40.7, 45.3) | 35.7 (32.7, 38.6) |
| Wait to see if safe then maybe later | 42.6 (35.6, 49.7) | 31.0 (28.6, 33.5) | 41.8 (38.6, 45.1) | 42.1 (39.8, 44.3) | 41.1 (38.0, 44.1) |
| Don't know if it will work | 28.2 (21.7, 34.8) | 23.9 (21.6, 26.2) | 22.9 (20.2, 25.6) | 25.9 (23.7, 28.0) | 23.6 (21.1, 26.1) |
| Allergic reaction | 19.6 (14.0, 25.2) | 20.5 (18.4, 22.6) | 26.8 (24.0, 29.7) | 26.4 (24.3, 28.6) | 23.2 (20.7, 25.8) |
| Don't like vaccines | 17.8 (11.7, 23.9) | 13.8 (11.9, 15.7) | 15.9 (13.5, 18.3) | 16.0 (13.9, 18.0) | 13.1 (11.1, 15.0) |
| Other people need more | 15.9 (11.1, 20.7) | 11.9 (10.3, 13.6) | 17.1 (14.3, 19.8) | 17.4 (15.3, 19.5) | 13.8 (11.7, 15.9) |
| Doctor not recommended | 12.0 (6.4, 17.6) | 8.3 (6.9, 9.6) | 10.0 (7.9, 12.2) | 9.3 (7.4, 11.2) | 10.0 (8.2, 11.8) |
| Safety concern because of health condition | 6.4 (3.2, 9.6) | 7.2 (6.0, 8.4) | 14.6 (12.3, 16.9) | 10.0 (8.2, 11.8) | 13.2 (11.2, 15.2) |
| Against religion | 13.0 (7.9, 18.1) | 7.9 (6.5, 9.3) | 9.5 (7.5, 11.5) | 7.6 (5.7, 9.4) | 9.3 (7.7, 11.0) |
| Currently/planning to be pregnant/breastfeeding | 6.6 (3.6, 9.6) | 2.5 (1.8, 3.3) | 15.9 (13.2, 18.5) | 9.6 (7.8, 11.5) | 13.3 (11.2, 15.4) |
| Cost | 2.1 (-0.2, 4.4) | 2.6 (1.7, 3.4) | 4.2 (2.7, 5.7) | 6.0 (4.2, 7.8) | 3.5 (2.4, 4.6) |
| Other | 22.6 (16.8, 28.3) | 15.2 (13.3, 17.2) | 10.2 (8.2, 12.2) | 10.9 (9.7, 12.1) | 13.6 (11.6, 15.6) |

[a]Including food processing, meat packing, laundry, and dry cleaning workers

[b]Not healthcare
